# Emotional dysregulation in childhood and disordered eating and self-harm in adolescence: Prospective associations and mediating pathways

**DOI:** 10.1101/2022.05.27.22275677

**Authors:** Naomi Warne, Jon Heron, Becky Mars, Francesca Solmi, Lucy Biddle, David Gunnell, Gemma Hammerton, Paul Moran, Marcus Munafò, Ian Penton-Voak, Andy Skinner, Anne Stewart, Helen Bould

**Author notes:** Correspondence to Dr Helen Bould, University of Bristol, Oakfield House, Oakfield Grove, Bristol, BS82BN.

## Abstract

**Background:** Emotional dysregulation may be a risk factor for disordered eating and self-harm in young people, but few prospective studies have assessed these associations long-term, or considered potential mediators. We examined prospective relationships between childhood emotional dysregulation and disordered eating and self-harm in adolescence; and social cognition, emotional recognition and being bullied as mediators.

**Methods:** We analysed Avon Longitudinal Study of Parents and Children data on 3453 males and 3481 females. We examined associations between emotional dysregulation at 7 years and any disordered eating and any self-harm at 16 years with probit regression models. We also assessed whether social cognition (7 years), emotional recognition (8 years) and bullying victimisation (11 years) mediated these relationships.

**Results:** Emotional dysregulation at age 7 years was associated with disordered eating (fully adjusted probit B (95% CI) = 0.082 (0.029, 0.134)) and self-harm (fully adjusted probit B (95% CI) = 0.093 (0.036, 0.150)) at age 16 years. There was no evidence of sex interactions or difference in effects between self-harm and disordered eating. Mediation models found social cognition was a key pathway to disordered eating (females 51.2%; males 27.0% of total effect) and self-harm (females 15.7%; males 10.8% of total effect). Bullying victimisation was an important pathway to disordered eating (females 17.1%; males 10.0% of total effect), but only to self-harm in females (15.7% of total effect). Indirect effects were stronger for disordered eating than self-harm.

**Conclusions:** In males and females, emotional dysregulation in early childhood is associated with disordered eating and self-harm in adolescence and may be a useful target for prevention and treatment. Mediating pathways appeared to differ by sex and outcome, but social cognition was a key mediating pathway for both disordered eating and self-harm.

## Introduction

Eating disorders affect up to 6% of female adolescents (Smink, Van Hoeken, Oldehinkel, & Hoek, 2014) and are associated with significant morbidity and mortality (Arcelus, Mitchell, Wales, & Nielsen, 2011). Disordered eating behaviours (including fasting, binge-eating and purging) are common (Warne et al., 2021), and associated with negative health and social outcomes, including eating disorders (Kärkkäinen, Mustelin, Raevuori, Kaprio, & Keski-Rahkonen, 2018; Tabler & Utz, 2015). Self-harm in young people is also common and associated with significant morbidity and mortality (Bergen et al., 2012; McManus et al., 2019). Critically, there is high comorbidity between disordered eating and self-harm. Prevalence of self-harm in patients with eating disorders is 14-64% (Cucchi et al., 2016; Svirko & Hawton, 2007); in the community, almost two-thirds of 16-year-old and 24-year-old females reporting self-harm also report disordered eating (Warne et al., 2021). This observed co-occurrence may reflect common aetiological pathways and shared risk factors (Svirko & Hawton, 2007).

Research indicates that emotional dysregulation - the inability to be aware of, accept, regulate, and modify emotional reactions and subsequent behaviours (Gratz & Roemer, 2004) - is linked to both eating disorders and self-harm. Emotional dysregulation is cross-sectionally associated with eating disorders (Lavender et al., 2015; Oldershaw, Lavender, Sallis, Stahl, & Schmidt, 2015) and self-harm (Colmenero-Navarrete, García-Sancho, & Salguero, 2021; Wolff et al., 2019). Some prospective studies have found associations between emotional dysregulation and eating disorders (Henderson et al., 2021; McLaughlin, Hatzenbuehler, Mennin, & Nolen-Hoeksema, 2011; Nolen-Hoeksema, Stice, Wade, & Bohon, 2007) and self-harm in adolescence (Garisch & Wilson, 2015; Robinson et al., 2019; Voon, Hasking, & Martin, 2014; You, Lin, & Leung, 2014; Zhou, Daukantaitė, Lundh, Wångby-Lundh, & Ryde, 2020). However, most of these studies follow-up for a year or less and focus on individual eating disorders/behaviours, or single maladaptive emotional regulation strategies, rather than global deficits. If prospective relationships for global emotional regulation deficits are established over longer periods, then it would enable more time to intervene on modifiable mediators, and potentially enable earlier interventions and effective prevention.

The mechanisms underlying the potential prospective associations of emotional dysregulation with disordered eating and self-harm are unknown. Three potential mediators of the association between emotional dysregulation and subsequent disordered eating and self-harm are social cognition, emotion recognition and bullying victimisation. Problems with social cognition (cognitive processes underlying social interaction such as perception, interpretation, and generation of responses to other people’s behaviours and mental states (Green et al., 2008)) in childhood are associated with higher subsequent rates of adolescent suicide attempts (Culpin et al., 2018), and disordered eating (Solmi et al., 2020). Cross-sectional studies find that people who self-harm (Seymour et al., 2016) and who have eating disorders (Caglar-Nazali et al., 2014) have increased difficulties with facial emotion recognition, and adolescents with dysregulated mood, a concept similar to emotional dysregulation, are less accurate than controls at identifying facial emotions (Rich et al., 2008). Emotional dysregulation is prospectively associated with bullying victimisation (Winsper, Hall, Strauss, & Wolke, 2017) and being bullied is associated with increased risk of subsequent self-harm (Bowes, Wolke, Joinson, Lereya, & Lewis, 2014; Lereya et al., 2013) and disordered eating (Copeland et al., 2015). Difficulties with social cognition and emotion recognition may therefore represent distinct mediating pathways from emotional dysregulation to eating disorders and/or self-harm, by, in turn, increasing a child’s susceptibility to bullying victimisation. Investigation of these factors as mediators has the potential to identify early modifiable targets and facilitate development of interventions to prevent both self-harm and eating disorders.

Our aims were to investigate the (1) association between emotional dysregulation in childhood and disordered eating and self-harm in adolescence in a population-based cohort and (2) extent to which social cognition, emotion recognition of others’ faces and being bullied mediate these associations.

## Methods

The research protocol was preregistered on Open Science Framework (https://osf.io/43kcg/). Deviations from the protocol are listed in Appendix S1.

### Study design and participants

We used data from a prospective birth cohort: the Avon Longitudinal Study of Parents and Children (ALSPAC) (Boyd et al., 2013; Fraser et al., 2013). Pregnant women with an expected delivery date between 1^st^ April 1991 and 31^st^ December 1992 living in the Avon area of Bristol, UK, were invited to take part. Of the 14,541 pregnancies, there were 13,988 children alive at 1 year of age. Ethical approval for the study was obtained from the ALSPAC Ethics and Law Committee and the Local Research Ethics Committees. Informed consent for the use of data collected via questionnaires and clinics was obtained from participants following the recommendations of the ALSPAC Ethics and Law Committee at the time. The study website contains details of all the available data through a fully searchable data dictionary and variable search tool: http://www.bristol.ac.uk/alspac/researchers/our-data/. We used a sample of 6,934 children (3,453 males, 3,481 females) from the core sample (Figure S1).

### Exposure

Mothers reported on their child’s emotional dysregulation (mean age 6 years 9 months) over the past 6 months using the Strengths and Difficulties Questionnaire-Dysregulation Profile (SDQ-DP) (Holtmann, Becker, Banaschewski, Rothenberger, & Roessner, 2011). This is a reliable, valid, and stable measure of emotional dysregulation (Deutz et al., 2018) composed of emotional, hyperactivity and conduct SDQ subscales (Table S1) (Goodman, 2001). To avoid small parameter estimates, and additional decimal places in quoted values, this variable was rescaled by dividing all values by 4, close to the standard deviation of the SDQ-DP in this sample (possible range 0-7.5). We chose not to standardize the variable as the SD would be expected to differ between complete case and imputed samples.

### Outcomes

Past-year self-harm and disordered eating were assessed via self-report questionnaire during adolescence (mean age 16 years 8 months).

#### Disordered eating

Questions were adapted from the Youth Risk Behaviour Surveillance System (Kann et al., 1996). Participants were asked about the presence and frequency of fasting (not eating for at least a day to lose weight/avoid gaining weight), purging (vomiting or taking laxatives/other medications to lose weight/avoid gaining weight), binge-eating (eating a large amount of food in a short amount of time, with a sense of loss of control), and excessive exercise (exercise in order to lose weight/avoid gaining weight that routinely interfered with daily routine or work) (Table S2). Our outcome was *any disordered eating* (fasting, purging, binge-eating, or excessive exercise at any frequency in the past year). Previous analysis of this data highlights that binge-eating and fasting are the more frequently reported behaviours (binge-eating: 16.2% of females, 4.3% of males; fasting: 20.7% of females, 3.4% of males), whereas smaller proportions of participants reported purging (9.5% of females, 1.9% of males) and excessive exercise (2.1% of females, 0.8% of males) (Warne et al., 2021).

#### Self-harm

We used two questions adapted from the Child and Adolescent Self-Harm in Europe study (Madge et al., 2008): whether the adolescent had hurt themselves on purpose (e.g., taking an overdose of pills or cutting themselves) and the last time they did so. Our outcome was *any self-harm* (regardless of suicidal intent) in the last year (Table S2) as suicidal intent was not measured.

### Mediators

Social cognition was measured using maternal reports on the Social and Communication Disorders Checklist (Skuse, Mandy, & Scourfield, 2005) at mean age 7 years 8 months (possible range 0-24). Emotion recognition was measured using the Diagnostic Assessment of Non-Verbal Accuracy (Nowicki & Duke, 1994) at mean age 8 years 8 months. We used the total number of facial emotions incorrectly identified (Reed et al., 2021) (possible range 0-24). For bullying victimisation, we used the summed score of bullying behaviours (Table S3) reported by participants at mean age 12 years 10 months on the Bullying and Friendship Interview Schedule (Wolke, Woods, Stanford, & Schulz, 2001) (possible range 0-27). Further details available in Appendix S2.

### Confounders

Potential confounders of the paths between exposure, mediators and outcomes included sex (recorded at birth), socioeconomic disadvantage, maternal mental health, and child general cognitive ability. We also considered Body Mass Index (BMI), at age 12 years 10 months, to be an intermediate confounder in mediation analyses. Details on confounder measurement and coding are available in Appendix S2.

### Statistical methods

We used a Structural Equation Modelling framework and the Weighted Least Squares estimator (WLSMV) in Mplus v8.3 (Muthén & Muthén, 2017).

#### Prospective associations

First, we used bivariate probit regression to determine associations between emotional dysregulation and the pair of correlated binary outcomes: disordered eating and self-harm. Wald tests were used to test whether 1) there was an overall association between emotional dysregulation and both outcomes, 2) the magnitude of the exposure-outcome effect differed between outcomes, and 3) interactions with sex. We sequentially adjusted for potential confounders. Probit coefficients can be interpreted as the effect that a one-unit increase in exposure has on the predicted z-score of the probability of the outcome. To aid interpretation, we derived marginal effects from the fully adjusted model and produced a margins plot showing how predicted probability of each outcome would be expected to vary across the observed range of emotional dysregulation.

#### Mediating pathways

Secondly, we extended the model, incorporating social cognition, emotion recognition and bullying victimisation as potential mediators (Figure 1). For this model, the sample was stratified to permit all paths to vary by sex. We adjusted all direct and indirect paths for confounders and additionally adjusted for the intermediate confounding effect of low or high BMI. As paths through BMI were not of substantive interest, indirect effect estimates for these specific paths were combined with those related to the three mediators of interest, or the direct effect, as appropriate (Figure S2).

**Figure 1.**
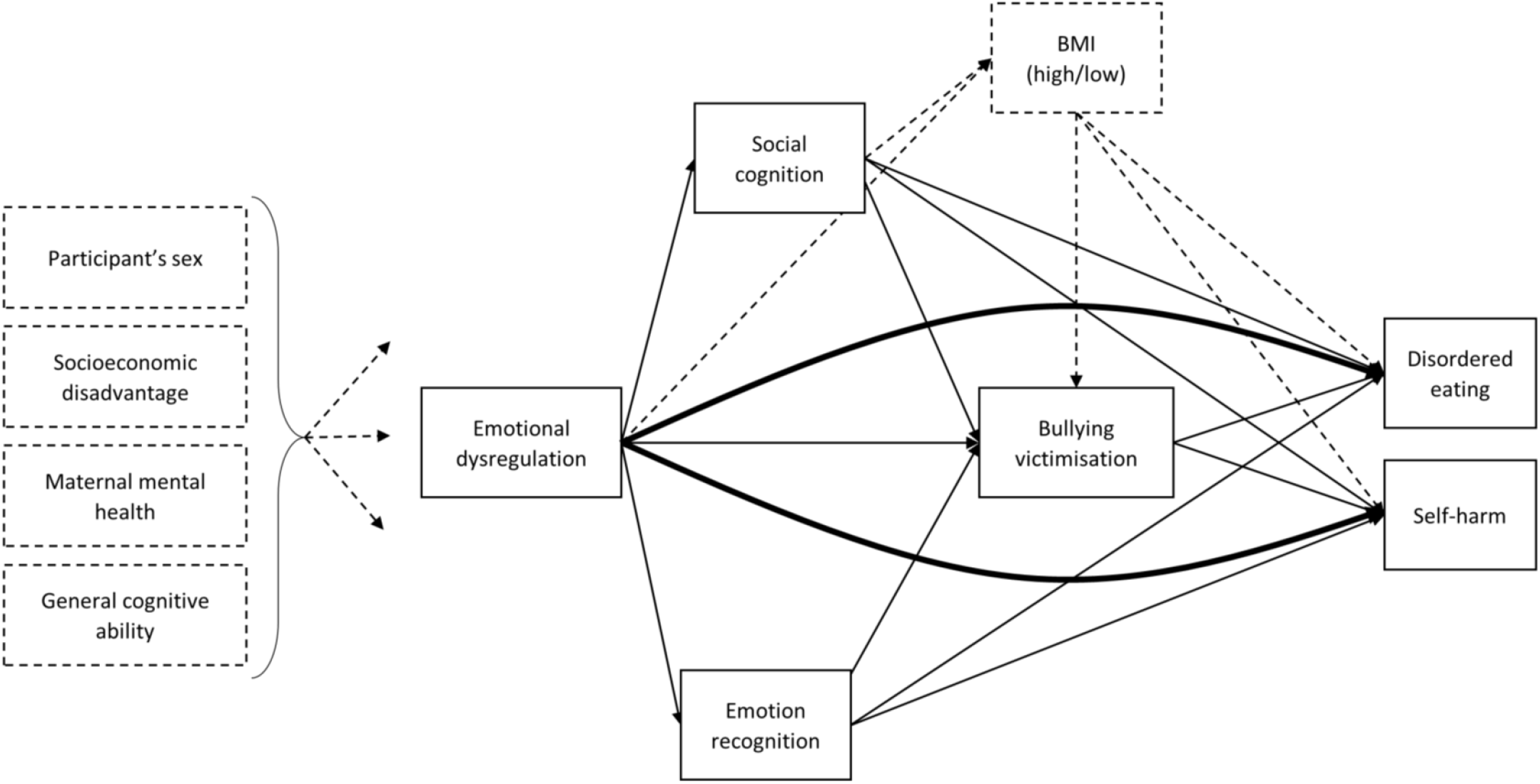
Hypothesised model linking emotional dysregulation with bivariate outcomes of self-harm and disordered eating Direct effects are depicted with lines of double thickness. Confounders and pathways involving confounders are depicted by dashed lines. For clarity, some paths between baseline confounders and mediators are not shown.

Indirect paths were quantified with the product-of-coefficients method, facilitated using probit regression for all binary/categorical dependent variables (MacKinnon, Lockwood, Brown, Wang, & Hoffman, 2007). We employed bootstrapping with 1000 bootstrap samples and the bootstrap SEs were used when pooling across imputed samples (“Rubin’s rules”). We express indirect effects as percentages of total effect. Finally, we explored associations for (a) each mediator/outcome pairing, and (b) the exposure and each mediator.

#### Missing data

We sought to address potential bias due to missing data by using a Fully Conditional Specification approach with -mi impute chained-in Stata version 16 (StataCorp, 2019). We used 25 cycles of regression-switching and produced 250 imputed datasets with imputation sample stratified by sex (more information in Appendix S3).

## Results

### Descriptive results

Descriptive statistics for the key variables are presented in Table 1. The sample had low levels of socioeconomic disadvantage and child ethnicity was predominantly white (95.9%) with a small proportion of children from non-white ethnic groups (4.1%). In the sample, maternal ethnicity (3.88% missing) was reported as white (94.4%), Black Caribbean (0.37%), Other Black (0.23%), Indian (0.37%), Pakistani (0.10%), Chinese (0.43%), and Other (0.43%). We were unable to report proportion of mothers reporting Black African ethnicity as ALSPAC stipulates censoring small cell sizes to preserve anonymity. At age 16, 15.3% females and 5.6% males reported past year self-harm, whereas 33.9% females and 9.4% males reported past year disordered eating. A comparison of observed and imputed samples for all model variables is available in Table S4.

**Table 1.**
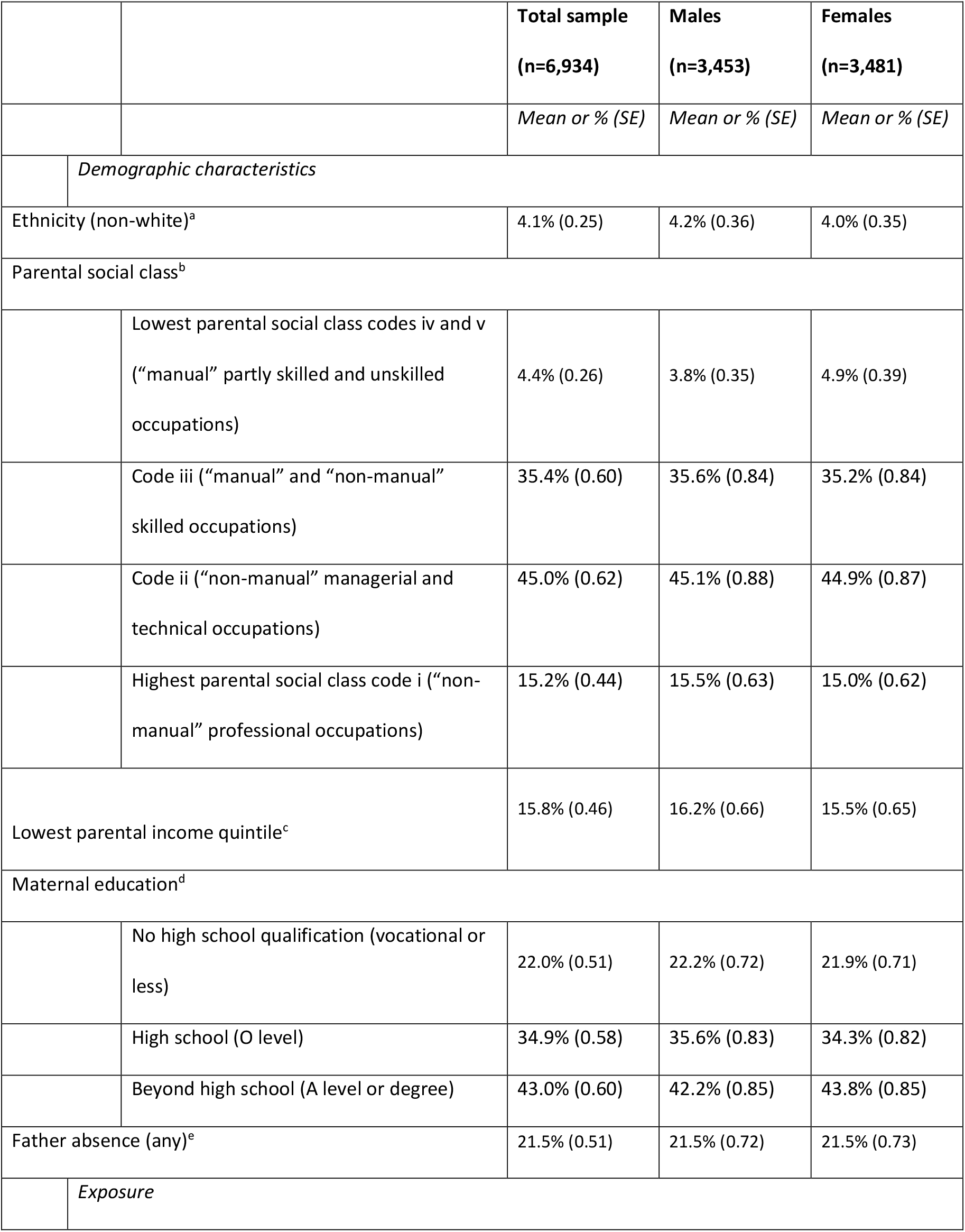
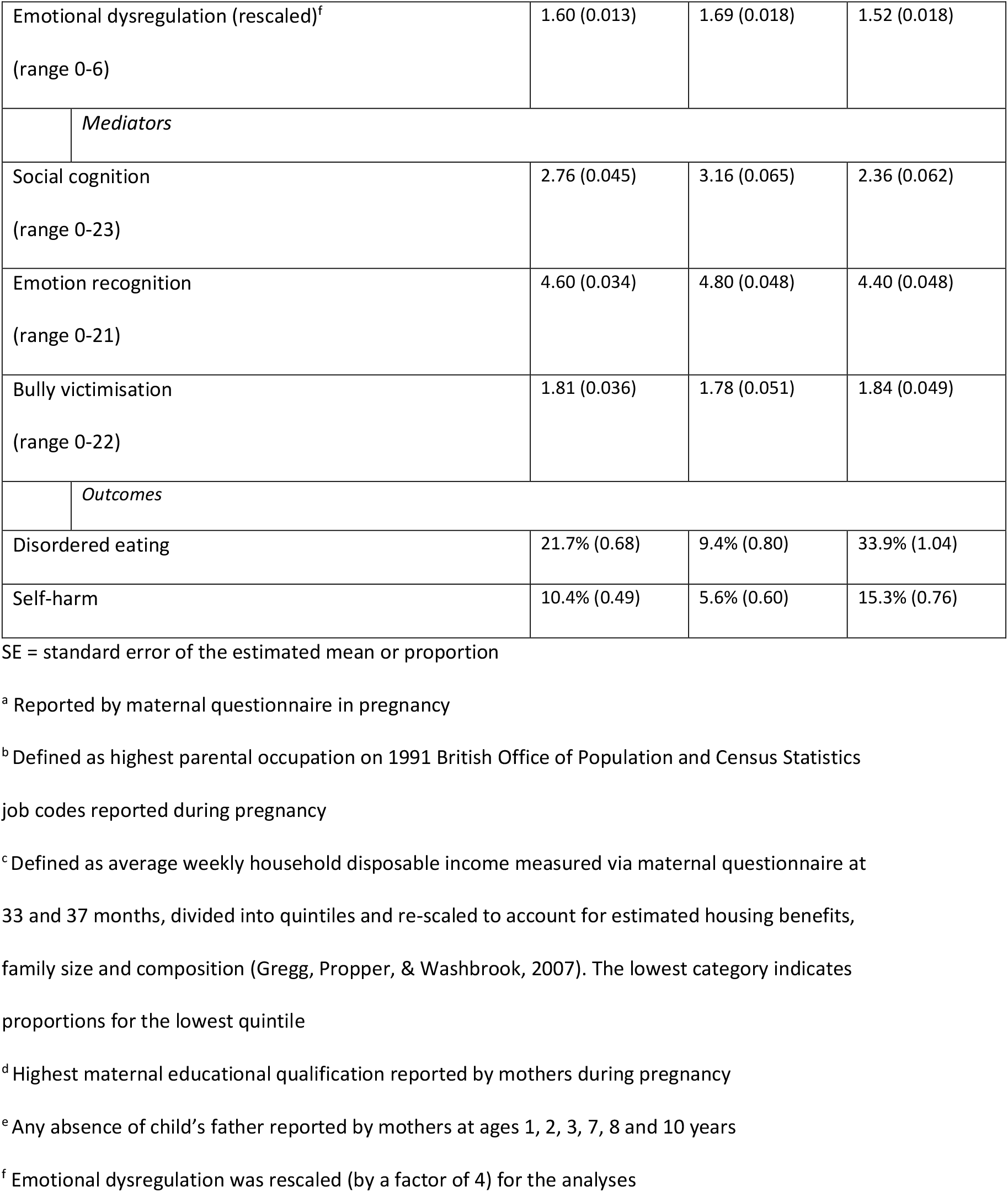
Descriptive statistics (imputed sample, n=6,934)

|  |  | **Total sample**<br>**(n=6,934)** | **Males**<br>**(n=3,453)** | **Females**<br>**(n=3,481)** |
| --- | --- | --- | --- | --- |
|  |  | *Mean or % (SE)* | *Mean or % (SE)* | *Mean or % (SE)* |
|  | *Demographic characteristics* |  |  |  |
| Ethnicity (non-white)<sup>a</sup> |  | 4.1% (0.25) | 4.2% (0.36) | 4.0% (0.35) |
| Parental social class<sup>b</sup> |  |  |  |  |
|  | Lowest parental social class codes iv and v<br>("manual" partly skilled and unskilled<br>occupations) | 4.4% (0.26) | 3.8% (0.35) | 4.9% (0.39) |
|  | Code iii ("manual" and "non-manual"<br>skilled occupations) | 35.4% (0.60) | 35.6% (0.84) | 35.2% (0.84) |
|  | Code ii ("non-manual" managerial and<br>technical occupations) | 45.0% (0.62) | 45.1% (0.88) | 44.9% (0.87) |
|  | Highest parental social class code i ("non-manual" professional occupations) | 15.2% (0.44) | 15.5% (0.63) | 15.0% (0.62) |
| Lowest parental income quintile<sup>c</sup> |  | 15.8% (0.46) | 16.2% (0.66) | 15.5% (0.65) |
| Maternal education<sup>d</sup> |  |  |  |  |
|  | No high school qualification (vocational or<br>less) | 22.0% (0.51) | 22.2% (0.72) | 21.9% (0.71) |
|  | High school (O level) | 34.9% (0.58) | 35.6% (0.83) | 34.3% (0.82) |
|  | Beyond high school (A level or degree) | 43.0% (0.60) | 42.2% (0.85) | 43.8% (0.85) |
| Father absence (any)<sup>e</sup> |  | 21.5% (0.51) | 21.5% (0.72) | 21.5% (0.73) |
|  | *Exposure* |  |  |  |

|  |  |  |  |
| --- | --- | --- | --- |
| Emotional dysregulation (rescaled)<sup>f</sup><br>(range 0-6) | 1.60 (0.013) | 1.69 (0.018) | 1.52 (0.018) |
| *Mediators* |  |  |  |
| Social cognition<br>(range 0-23) | 2.76 (0.045) | 3.16 (0.065) | 2.36 (0.062) |
| Emotion recognition<br>(range 0-21) | 4.60 (0.034) | 4.80 (0.048) | 4.40 (0.048) |
| Bully victimisation<br>(range 0-22) | 1.81 (0.036) | 1.78 (0.051) | 1.84 (0.049) |
| *Outcomes* |  |  |  |
| Disordered eating | 21.7% (0.68) | 9.4% (0.80) | 33.9% (1.04) |
| Self-harm | 10.4% (0.49) | 5.6% (0.60) | 15.3% (0.76) |

SE = standard error of the estimated mean or proportion
<sup>a</sup> Reported by maternal questionnaire in pregnancy
<sup>b</sup> Defined as highest parental occupation on 1991 British Office of Population and Census Statistics job codes reported during pregnancy
<sup>c</sup> Defined as average weekly household disposable income measured via maternal questionnaire at 33 and 37 months, divided into quintiles and re-scaled to account for estimated housing benefits, family size and composition (Gregg, Propper, & Washbrook, 2007). The lowest category indicates proportions for the lowest quintile
<sup>d</sup> Highest maternal educational qualification reported by mothers during pregnancy
<sup>e</sup> Any absence of child's father reported by mothers at ages 1, 2, 3, 7, 8 and 10 years
<sup>f</sup> Emotional dysregulation was rescaled (by a factor of 4) for the analyses

### Prospective associations

Table 2 shows probit estimates for the imputed sample. There was modest evidence (p=0.019) for a positive association between emotional dysregulation and both outcomes, however estimates increased markedly in magnitude following adjustment for sex. In the fully adjusted model, a one unit increase in our emotional dysregulation scale was associated with a 0.082 (95% CI 0.029, 0.134) increase in z-score for probability of disordered eating and a 0.093 (95% CI 0.036, 0.150) increase in z-score for probability of self-harm (Figure 2). As shown in Table S5, there was little evidence that emotional dysregulation had a differential impact on disordered eating and self-harm (difference in estimates p=0.745) and little evidence that the magnitude of associations varied by sex (interaction p=0.600). Complete case sample results were consistent with imputed sample results (Table S5).

**Table 2.** Prospective association between childhood emotional dysregulation and adolescent disordered eating and self-harm (imputed sample, n=6,934)

|  | Outcome |  | Wald test (2 d.f.), p-value |
| --- | --- | --- | --- |
|  | Disordered eating<br>Probit B (95% CI) | Self-harm<br>Probit B (95% CI) |  |
| Unadjusted | 0.048 (0.003, 0.094) | 0.061 (0.011, 0.111) | 7.91, 0.0192 |
| Adjusted for sex | 0.095 (0.045, 0.145) | 0.086 (0.034, 0.138) | 20.10, <0.001 |
| Additionally adjusted for socioeconomic disadvantage | 0.086 (0.036, 0.136) | 0.086 (0.033, 0.140) | 17.57, <0.001 |
| Additionally adjusted for maternal mental health | 0.076 (0.024, 0.127) | 0.075 (0.019, 0.131) | 12.47, 0.002 |
| Fully adjusted for all baseline confounders | 0.082 (0.029, 0.134) | 0.093 (0.036, 0.150) | 15.95, <0.001 |

Wald test = Omnibus test of the association between exposure and both outcomes

**Figure 2.**
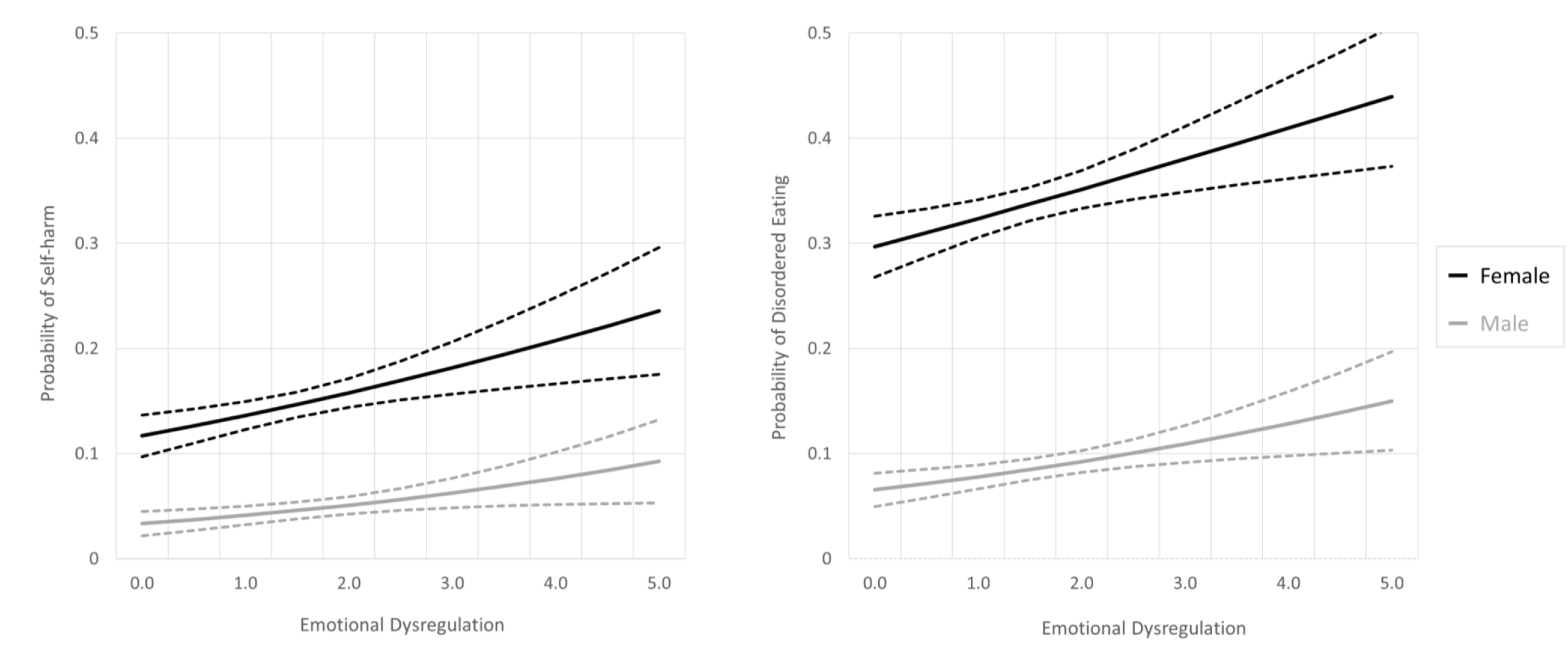
The predicted probability of self-harm and disordered eating across the range of emotional dysregulation. All baseline confounders, other than sex, have been held at their mean levels. Dotted lines indicate 95% CI.

### Mediating pathways

Estimated indirect effects (Table 3) show differences in the proportion of the total effect mediated for each combination of sex and outcome which ranged from 15.7% total indirect effect for self-harm in males, to 62.2% for disordered eating in females.

**Table 3.** Mediating pathways from emotional dysregulation to disordered eating and self-harm (imputed sample, n=6,934)

|  | Males |  | Females |  |
| --- | --- | --- | --- | --- |
|  | Disordered eating | Self-harm | Disordered eating | Self-harm |
|  | Probit B (SE)<br>% of<br>total | Probit B (SE)<br>% of<br>total | Probit B (SE)<br>% of<br>total | Probit B (SE)<br>% of<br>total |
| Total effect | 0.100 (0.053) | 0.102 (0.055) | 0.082 (0.034) | 0.096 (0.036) |
| Direct effect | 0.076 (0.063) 76.0% | 0.085 (0.065) 83.3% | 0.030 (0.039) 36.6% | 0.069 (0.042) 71.9% |
| Total indirect effect | 0.024 (0.034) 24.0% | 0.016 (0.034) 15.7% | 0.051 (0.020) 62.2% | 0.027 (0.022) 28.1% |
| Emotional dysregulation → social cognition → outcome | 0.021 (0.033) 21.0% | 0.009 (0.033) 8.8% | 0.038 (0.020) 46.3% | 0.011 (0.022) 11.5% |
| Emotional dysregulation → emotion recognition → outcome | -0.007 (0.004) -7.0% | 0.003 (0.004) 2.9% | -0.001 (0.001) -1.2% | 0.001 (0.002) 1.0% |
| Emotional dysregulation → bullying victimisation → outcome | 0.004 (0.003) 4.0% | 0.002 (0.003) 2.0% | 0.010 (0.004) 12.2% | 0.011 (0.005) 11.5% |
| Emotional dysregulation → social cognition → bullying victimisation → outcome | 0.005 (0.004) 6.0% | 0.002 (0.004) 2.0% | 0.004 (0.002) 4.9% | 0.004 (0.002) 4.2% |
| Emotional dysregulation → emotion recognition → bullying victimisation → outcome | 0.000 (0.000) 0.0% | 0.000 (0.000) 0.0% | 0.000 (0.000) 0% | 0.000 (0.000) 0.0% |

% of total = percentage of the total effect explained by the pathway. Fully adjusted for all confounders. SE incorporates bootstrapping (1000 iterations) and pooling across 250 imputed datasets.

Social cognition was the dominant mediator between emotional dysregulation and both self-harm and disordered eating, with substantial pathways via social cognition alone, and via both social cognition and bullying. In females, social cognition accounted for 51.2% (46.3%+4.9%) of the total effect of emotional dysregulation on disordered eating, and 15.7% (11.5%+4.2%) for self-harm; whilst in males these figures were lower, at 27.0% (21.0%+6.0%) and 10.8% (8.8%+2.0%) respectively.

In females, mediation via bullying victimisation accounted for 17.1% (12.2%+4.9%+0.0%) of the total effect on disordered eating and 15.7% (11.5%+4.2%+0.0%) of the total effect on self-harm. For males, bullying victimisation accounted for 10.0% (6.0%+4.0%+0.0%) of the total effect on disordered eating but only 4.0% (2.0%+2.0%+0.0%) of the total effect for self-harm.

Finally, emotion recognition accounted for a negligible proportion of the total effect for both outcomes in both females and males. Additional analyses (Tables S6 and S7) found that whilst emotion recognition was strongly related to emotional dysregulation (particularly for males) the association between emotion recognition and both self-harm and disordered eating was modest.

## Discussion

Children with higher emotional dysregulation were more likely to report disordered eating and self-harm during adolescence. Mediating pathways appeared to differ by sex (females had two dominant indirect effects via social cognition and bullying, whereas males had these two dominant pathways for disordered eating, but only one dominant indirect effect (social cognition) for self-harm). Pathways also differed by outcome, with stronger indirect effects found for disordered eating than self-harm. These results suggest that emotional dysregulation, social cognition, and potentially bullying, may be useful targets to prevent disordered eating and self-harm.

Our findings are consistent with previous literature which has found positive prospective associations between emotional dysregulation and self-harm (Garisch & Wilson, 2015; Robinson et al., 2019; Voon et al., 2014; You et al., 2014; Zhou et al., 2020) and disordered eating (Henderson et al., 2021; McLaughlin et al., 2011; Nolen-Hoeksema et al., 2007). Our study extends these findings to a longer follow up (9 years) from childhood to adolescence, and considers both self-harm and disordered eating, which may share some aetiological factors (Svirko & Hawton, 2007). In combination with previous literature, our findings suggest that interventions targeting emotional dysregulation may be useful for prevention and treatment of eating disorders and self-harm, as it is modifiable at the individual level (Gratz, Weiss, & Tull, 2015). Improving emotional regulation skills may be useful as a universal transdiagnostic preventative intervention for adolescents (Volkaert, Wante, Loeys, Boelens, & Braet, 2021). However, further longitudinal research is needed to understand potential preventative effects for disordered eating and self-harm. Training in emotional regulation skills can be helpful for adults with eating disorders (Davies et al., 2012) and self-harm (DeCou, Comtois, & Landes, 2019) and preliminary evidence suggests it may also be useful in adolescents (Giombini et al., 2021; Kothgassner et al., 2021).

Social cognition was an important mediating pathway for both disordered eating and self-harm, consistent with literature on prospective links between autism spectrum traits and disordered eating (Solmi et al., 2020) and self-harm (Culpin et al., 2018). Our work extends previous findings and suggests that emotional dysregulation may precede these associations. However, we cannot rule out the possibility that social cognition problems measured at age 7 represent earlier differences in social cognition that precede emotional dysregulation. One way to sever the link between emotional dysregulation and subsequent disordered eating and self-harm could be to develop interventions that de-stigmatise atypical social cognition in children. Alternatively, interventions to strengthen social cognition (Fernández-Sotos et al., 2018), or strongly related concepts such as mentalisation (the capacity to interpret and understand behaviour from oneself and others based on underlying mental states (Simonsen, Jakobsen, Grøntved, & Kjaersdam Telléus, 2020)) may have a place in the treatment of eating disorders and self-harm. Previous research has found lower mentalisation ability is associated with eating disorders (Jewell et al., 2016; Simonsen et al., 2020), deficits in mentalisation (excessive certainty about mental states) predict poorer treatment outcomes for adolescents with anorexia nervosa (Jewell et al., 2021). Mentalisation-based therapies may also be effective in improving core body image psychopathology in adults with eating disorders (Robinson et al., 2016), and reducing self-harm in adolescents (Griffiths et al., 2019; Rossouw & Fonagy, 2012). We found stronger indirect effects in relation to disordered eating than for self-harm, suggesting that addressing difficulties with social cognition or mentalisation may be more effective in reducing or preventing disordered eating than self-harm. Further research on associations between difficulties in social cognition, mentalisation and eating patterns is warranted.

Bullying victimisation was also a key mediating pathway for disordered eating and self-harm in females, but only for disordered eating in males. The lack of association in males may have been driven by very weak associations between bullying victimisation and self-harm (Table S6), and it is possible that bullying victimisation led to alternative negative outcomes in males that were not assessed. Nevertheless, our findings suggest that effective anti-bullying interventions (e.g. in schools (Fraguas et al., 2021)) may be beneficial in preventing disordered eating regardless of sex, and in preventing self-harm in females.

In the context of our analysis, emotion recognition was not an important mediator. Emotional dysregulation was associated with emotion recognition, but we did not find strong associations between emotion recognition and either disordered eating or self-harm. It may be that difficulties in recognising specific emotions (e.g. anger and fear) are more important than overall accuracy in emotion recognition. It is also important to acknowledge that our emotion recognition task focused on facial emotions of others, but difficulties associated with recognising one’s own emotions (i.e. alexithymia) may help explain the relationship between emotion dysregulation and subsequent disordered eating and self-harm. Indeed associations have previously been reported between alexithymia and eating disorders, symptoms of eating disorders (Nowakowski, McFarlane, & Cassin, 2013; Shank et al., 2019) and also with self-harm (Norman, Oskis, Marzano, & Coulson, 2020). We did not have the data to explore this in the current study, but future research would benefit from considering recognition of one’s own emotions.

### Strengths and limitations

Strengths include using a large prospective cohort with considerable information on potential confounders and including both self-harm and disordered eating. Limitations include measuring any frequency of disordered eating rather than diagnostic-level frequency or a research diagnosis of eating disorders. This was so we could assess the relationship between emotional dysregulation across a spectrum of potentially impairing eating behaviours that are not always picked up in clinical samples. Our results may not generalise to individuals with eating disorder diagnoses. We were unable to differentiate self-harm with and without suicidal intent in the last year as this data was not available at age 16. The sample was mainly white and less socioeconomically disadvantaged, so results may not generalise to other ethnic and socioeconomic groups. Furthermore, we were unable to list all of the races and ethnicities present in the sample (i.e. list the “non-white” child ethnicity groups separately) as, due to small numbers, this data was censored to preserve anonymity. Findings on social cognition as a potential mediator should be interpreted cautiously as social cognition was measured close in time to the exposure, and by the same informant, which may have inflated associations. The measure of bullying did not include items specifically pertaining to bullying in relation to appearance or weight, so the strength of association of bullying in relation to subsequent disordered eating in both boys and girls may be an underestimate. Although we adjusted for several important confounders, it is possible there may be residual confounding. Another limitation is that although we measured our putative mediators at a later time point than emotional dysregulation, this does not mean that deficits in social cognition and emotion recognition are necessarily a consequence of emotional dysregulation. Indeed, it is possible that that difficulties with emotion regulation, social cognition, and emotion recognition are all indicators of a broader deficit in social and emotional processing, rather than causal of each other. Further work should determine whether these risk factors represent distinct phenomena. Finally, we did not statistically examine the difference in proportion-mediated for males and females. Consequently, sex differences should be interpreted with some caution.

## Conclusion

Emotional dysregulation in childhood may be an important precursor for adolescent disordered eating and self-harm. Social cognition was a key mediating pathway for both outcomes and sexes and being bullied was important for disordered eating in both males and females, and self-harm in females. Indirect effects were stronger for disordered eating than for self-harm. Findings suggest that emotional dysregulation, social cognition and bullying may be useful targets in the prevention and treatment of disordered eating and self-harm.

## Key points and relevance

Key points and relevance

- Few studies have examined associations between childhood emotional dysregulation and adolescent disordered eating/self-harm
- We found emotional dysregulation (7 years) was associated with both self-harm and disordered eating (16 years), but associations did not differ by sex or outcome
- Measuring emotional dysregulation may be a useful way of identifying young people at high risk of developing disordered eating/self-harm and a potential target for interventions
- Social cognition and bullying were important mediating pathways and may be targets for interventions

## Data Availability

ALSPAC data access is through a system of managed open access. The steps below highlight how to apply for access to the data included in this paper and all other ALSPAC data. 1. Please read the ALSPAC access policy (http://www.bristol.ac.uk/media-library/sites/alspac/documents/researchers/data-access/ALSPAC_Access_Policy.pdf) which describes the process of accessing the data and samples in detail, and outlines the costs associated with doing so. 2. You may also find it useful to browse our fully searchable research proposals database, which lists all research projects that have been approved since April 2011. 3. Please submit your research proposal (https://proposals.epi.bristol.ac.uk/) for consideration by the ALSPAC Executive Committee. You will receive a response within 10 working days to advise you whether your proposal has been approved. If you have any questions about accessing data or samples, please (data) or bbl-info{at}bristol.ac.uk (samples).

## Abbreviations

ALSPAC: Avon Longitudinal Study of Parents and Children

## Acknowledgements

We are extremely grateful to all the families who took part in this study, the midwives for their help in recruiting them, and the whole ALSPAC team, which includes interviewers, computer and laboratory technicians, clerical workers, research scientists, volunteers, managers, receptionists, and nurses.

