## Appendix for "Emotional dysregulation in childhood and disordered eating and self-harm in adolescence: Prospective associations and mediating pathways"

### Supplementary Information

#### Appendix S1 Deviations from published protocol

Prior to starting analysis we published an analysis plan on the Open Science Framework: https://osf.io/43kcg/

In this we aimed to investigate outcomes at age 16 and age 24 years. However in the final manuscript we limit analyses to age 16 only. Whilst we could have conducted multivariable regression at age 24 years, we could not conduct an appropriate mediation model with age 24 outcomes. We considered two approaches: 1) keeping all variables the same in a mediation model but including outcomes at age 24 instead of age 16 years, and 2) including relevant mediators at later time points in a new mediation model with outcomes at 24 years. However, when mapping both of these approaches, data availability precluded robust analyses accounting for important cofounders. With the first approach we would be missing key mediators and confounders on the pathway from the latest mediator (bullying victimisation at 12 years) and outcomes at age 24 years, and with the second approach there would be issues with reverse causality as hypothesised mediators were only available after peak onset of outcomes. Therefore, we did not perform analysis using outcomes at age 24 years; this decision was made prior to the start of analysis.

We also included paternal absence as an additional indicator of socioeconomic disadvantage in the baseline confounders.

**Appendix S2. Information on mediators and confounders**

***Social cognition (mediator)***

The Social and Communication Disorders Checklist (Skuse, Mandy, & Scourfield, 2005) comprises 12 statements with responses of “not true” (0), “quite true or sometimes true” (1), and “very or often true” (2) over the past 6 months. The summed score (range 0-24) has been shown to have high reliability (Skuse et al., 2005) and is a strong predictor of Autism Spectrum Disorders in the ALSPAC cohort (Rai et al., 2018).

***Emotion recognition (mediator)***

Emotion recognition was measured using the Diagnostic Assessment of Non-Verbal Accuracy (Nowicki & Duke, 1994), which includes a sub-test of ability to recognise facial displays of emotion (“DANVA faces”). This was administered during a face-to-face clinic at mean age 8 years 8 months. Participants were asked to identify the emotion displayed (high and low intensity for happy, sad, angry, or afraid) in 24 photographs of child faces, each displayed for two seconds. We used the total number of facial emotions incorrectly identified (Reed et al., 2021).

***Bullying victimisation (mediator)***

At mean age 12 years 10 months, participants indicated the presence and frequency of nine (overt and relational) bullying behaviours experienced over the past 6 months on the Bullying and Friendship Interview Schedule (Wolke, Woods, Stanford, & Schulz, 2001) (Table S3). Item responses were rated on a scale of “never” (0), “seldom (<4 times)” (1), “frequently (≥4 times)” (2), or “very frequently (at least once a week)” (3). We summed responses to create a continuous score (range 0-27) (Bowes, Joinson, Wolke, & Lewis, 2015).

***Socioeconomic disadvantage (confounders)***

A number of indicators of socioeconomic disadvantage were used as confounders due to associations with emotional dysregulation (Chen & Miller, 2014; Troy, Ford, McRae, Zarolia, & Mauss, 2017), eating disorders (Mitchison, Hay, Slewa-Younan, & Mond, 2014), and self-harm (Hu, Li, Glauert, & Taylor, 2017; Lodebo, Möller, Larsson, & Engström, 2017).

Indicators of socioeconomic disadvantage included highest paternal or maternal social class (reported during pregnancy); maternal education (reported at 32 weeks’ gestation as mother’s highest educational qualification (coded as no high school qualifications, high school, beyond high school); average weekly income (quintiles measured on maternal questionnaires at 33 and 47 months); paternal absence (coded no paternal absence, paternal absence when child was 5 years or older, paternal absence when child was less than 5 years old); and maternal age at delivery.

***Maternal mental health (confounders)***

Maternal mental health may affect child's emotional dysregulation (Stein et al., 2014) and is also linked to offspring eating disorder (Bould et al., 2015) and self-harm (Hu et al., 2017). Maternal depressive symptoms were reported on the Edinburgh Postnatal Depression Scale (EPDS) (Cox, Holden, & Sagovsky, 1987) at 32 weeks of gestation and at 21 months post-partum.

Maternal lifetime eating disorder (self-reported bulimia and/or anorexia nervosa) was assessed in pregnancy (12 weeks gestation). Mothers were asked "have you ever had any of the following problems?" to a number of physical and mental health issues including bulimia and anorexia. Mothers who reported "yes had it recently" or "yes in past, not now" to bulimia and anorexia items were considered to have a lifetime eating disorder.

***General cognitive ability (confounder)***

General cognitive ability may be linked to later self-harm (Chang et al., 2014) and emotion recognition (Andric, Maric, Mihaljevic, Mirjanic, & van Os, 2016; Buitelaar, Wees, Swaab-Barneveld, & Gaag, 1999), and is plausibly associated with emotional dysregulation and social cognition. Full scale IQ was assessed at age 8 years in a face-to-face clinic session using the Wechsler Intelligence Scale for Children-III (WISC-III) (Wechsler, Golombok, & Rust, 1992).

***Body mass index (BMI)***

BMI has a clear association with disordered eating (APA, 2013). However, evidence of a link between BMI and self-harm is mixed, ranging from no link between BMI and self-harm (Mewton et al., 2019), to associations with low BMI (Kinoshita et al., 2012), both low and high BMI (Nyberg, Gustavsson, Åberg, Kuhn, & Waern, 2019), and associations for high BMI and suicide (Amiri & Behnezhad, 2018; Magnusson, Rasmussen, Lawlor, Tynelius, & Gunnell, 2006). Evidence also suggests higher BMI may be linked to emotional dysregulation (Anderson, Sacker, Whitaker, & Kelly, 2017; Gianini, White, & Masheb, 2013; Sainsbury et al., 2019), social cognition (Ly et al., unpublished) and being bullied (Hammami, Chaurasia, Bigelow, & Leatherdale, 2020; Janssen, Craig, Boyce, & Pickett, 2004).

We considered BMI to be an intermediate confounder that potentially confounds the bullying to self-harm/disordered eating relationships but is causally downstream of both Emotional Dysregulation and Social Cognition (see Figure 1). Therefore, we used BMI derived from height and weight measurements taken by ALSPAC researchers (mean age 12 years, 10 months). We used the Stata zbmicat function (Vidmar, Carlin, Hesketh, & Cole, 2013) to generate age- and sex-appropriate BMI categories of underweight (BMI $<18.5$), normal weight (BMI 18.5 to $<25$) and overweight (BMI $\geq 25+$). In analyses we use two binary variables and refer to these as low BMI (versus high and normal BMI) and high BMI (versus low and normal BMI).

**Appendix S3 Approach to missing data**

We sought to address the potential problem of bias due to missing data by using Fully Conditional Specification (FCS) approach with -mi impute chained- in Stata version 16 (StataCorp, 2019). We used 25 cycles of regression-switching and produced 250 imputed datasets with the imputation sample stratified by sex using the -by()- option to enable gender to moderate all associations. Incomplete binary variables were imputed using logistic regression, with ordinal and multinomial regression employed for category variables. Depending on variable distributions we imputed continuous data either under a normal model or using prediction mean matching with ten nearest-neighbours. Finally, auxiliary data were introduced to improve the validity of the required Missing At Random (MAR) assumption. In most cases the specific auxiliary variables varied from one prediction equation to the next and consisted of strong correlates of the incomplete variable being imputed, such as more-complete measurements collected at previous data-collection waves. The imputation sample was restricted to 6,934 participants (3,453 males and 3,481 females) due to a lack of good auxiliary data for the incomplete measure of IQ from age 8. Thus our chosen strategy was based on the assumption that IQ was likely to be Missing Not At Random (MNAR), however within the sample of cases with complete IQ, the other missing data would be MAR and hence imputable using the observed data to hand. Supplementary Table S4 shows rates or means for each imputation variable from the substantive model and how these varied between the observed cases and those which were imputed. For the purpose of this summary table, all categorical confounders were dichotomised to simplify the output.

*Reflection on limitations of our chosen approach*

We employed multiple imputation to handle partial missingness within a subset of the ALSPAC sample, thus boosting our analysis-sample to approximately half of the original cohort. We were hampered by a lack of auxiliary information with which to impute IQ, so this variable is likely to be Missing Not at Random (MNAR) and therefore, as far as IQ is concerned, the subsample defined by missing(IQ)=0 may not be representative of the wider ALSPAC study sample. Recent work (Hughes, Heron, Sterne, & Tilling, 2019) has shown that with bias due to missing data, the crucial aspect is whether there are conditional differences in the outcome distributions between those included and excluded from the analysis. It follows from this work that provided our outcomes are conditionally unrelated (i.e. conditional on the independent variables from our model) to whether the children attended a face-to-face clinic at age 8 year, then our findings should be unbiased. Given the range of confounders considered here, we believe this assumption is reasonable.

**Appendix S4. Additional funding**

Becky Mars is funded by a Medical Research Foundation Fellowship (MRF-058-0017-F-MARS-C0869, MRF-058-0017-F-MARS-C0869s1).

David Gunnell, Ian Penton-Voak and Paul Moran are part-funded by the NIHR Biomedical Research Centre at University Hospitals Bristol and Weston NHS Foundation Trust and the University of Bristol.

Francesca Solmi and Gemma Hammerton are funded by Sir Henry Wellcome Fellowships (209196/Z/17/Z and 209138/Z/17/Z).

Paul Moran and Lucy Biddle are part-funded by NIHR Applied Research Collaboration (ARC) West.

Marcus Munafò is a member of the MRC Integrative Epidemiology Unit at the University of Bristol (MC_UU_00011/7).

Andy Skinner is supported by Cancer Research UK (C18281/A19169, C18281/A29019) and the MRC Integrative Epidemiology Unit at the University of Bristol (R120971-106).

The views expressed in this correspondence are those of the authors and not necessarily those of the National Health Service, the National Institute for Health Research, or the Department of Health and Social Care.

**Table S1. Strengths and Difficulties Questionnaire – Dysregulation Profile (SDQ-DP) items**

| Questionnaire Item | Response options |
|---|---|
| In the last 6 months she/he…. | |
| **Emotional problems** | 0 – Not true<br>1 – Somewhat true<br>2 – Certainly true |
| 3. Often complained of headaches, stomach-aches or sickness | |
| 8. Has many worries, often seems worried | |
| 13. Often unhappy, down-hearted or tearful | |
| 16. Is nervous or clingy in new situations, easily loses confidence | |
| 24. Has many fears, is easily scared | |
| **Conduct problems** | |
| 5. Often has temper tantrums or hot tempers | |
| 7. Is generally obedient, usually does what adults request (R) | |
| 12. Often fights with other children or bullies them | |
| 18. Often lies or cheats | |
| 22. She/he steals from home, school, or elsewhere | |
| **Hyperactivity-inattention** | |
| 2. Has been restless, overactive, cannot stay still for long | |
| 10. Constantly fidgeting or squirming | |
| 15. Easily distracted, concentration wanders | |
| 21. Thinks things out before acting (R) | |
| 25. Sees tasks through to the end, has good attention span (R) | |

(R) = reverse coded

Values shown in first column reflect the order the questions were asked as part of the broader SDQ battery.

**Table S2. Disordered eating and self-harm questions and variable derivation at 16 years in ALSPAC**

Any Disordered Eating = Any Fasting OR Any Purging OR Any Excessive Exercise OR Any Binge-eating

| Questions | Response options | Coding | Final variable |
|---|---|---|---|
| **Fasting** | | | |
| During the **past year**, how often did you fast (not eat for at least a day) to lose weight or avoid gaining weight? | 1. Never<br>2. Less than once a month<br>3. 1-3 times a month<br>4. Once a week<br>5. More than once a week | Any fasting<br>1 = "no"<br>2-5 = "yes" | **Any fasting** |
| **Purging** | | | |
| During the **past year**, how often did you make yourself throw up (vomit) to lose weight or avoid gaining weight? | 1. Never<br>2. Less than once a month<br>3. 1-3 times a month<br>4. Once a week<br>5. 2-6 times a week<br>6. Every day | Any self-induced vomiting<br>1 = "no"<br>2-6 = "yes" | **Any purging**<br><br>Any self-induced vomiting<br>OR<br>Any laxative/medication use |
| a) During the **past year**, did you take laxatives or other tablets or medicines (diet pills or water tablets) to lose weight or avoid gaining weight?<br><br>b) How often? | 1. Yes, laxative<br>2. Yes, other<br>3. Never<br><br>1. Never<br>2. Less than once a month<br>3. 1-3 times a month<br>4. Once a week<br>5. 2-6 times a week<br>6. Every day | Any laxative/medication use<br>a) 3 = "no"<br>a) 1-2 = "yes"<br>OR<br>b) 2-6 = "yes" | |
| **Excessive exercise** | | | |
| During the **past year**, how often did you do any exercise (going to the gym, brisk walking or any sports activity)? | 1. 5 or more times a week<br>2. 1-4 times a week<br>3. 1-3 times a month<br>4. Less than once a month<br>5. Never | Any exercise<br>5 = "no"<br>1-4 = "yes" | **Any excessive exercise**<br><br>Any exercise to lose weight<br>AND<br>Exercise interfered with life<br>(= 0 if Any exercise = "no") |
| Did you exercise in order to lose weight or avoid gaining weight? | 1. Yes, sometimes<br>2. Yes, frequently<br>3. No | Any exercise to lose weight<br>3 = "no"<br>1-2 = "yes" | |
| Was it difficult for you to do your work or schoolwork because of the amount of time that you were exercising? | 1. Yes, sometimes<br>2. Yes, frequently<br>3. No | Exercise interfered with life<br>1 & 3 = "no"<br>2 = "yes" | |
| **Binge eating** | | | |
| Sometimes people will go on an 'eating binge', where they eat an amount of food that most people would consider to be **very** large, **in a short period of time.** During the **past year**, how often did you go on an eating binge? | 1. Less than once a month<br>2. 1-3 times a month<br>3. Once a week<br>4. More than once a week<br>5. Never | Any bingeing<br>5 = "no"<br>1-4 = "yes" | **Any binge-eating**<br><br>Any bingeing<br>AND<br>Loss of control |
| Did you feel out of control, like you couldn't stop eating even if you wanted to? | 1. Yes, usually<br>2. Yes, sometimes<br>3. No | Loss of control<br>3 = "no"<br>1-2 = "yes" | |

**Table S2 (cont.) Disordered eating and self-harm questions and variable derivation at 16 years in ALSPAC**

| Questions | Response options | Coding | Final variable |
|---|---|---|---|
| **Self-harm** | | | |
| When was the **last time** you hurt yourself on purpose? | 1. In the last week<br>2. More than a week ago but in the last year<br>3. More than a year ago | Self-harm past year<br>3 = "no"<br>1-2 = "yes" | **Self-harm**<br><br>Self-harm past year<br>(= 0 if self-harm ever = no) |
| Have you ever hurt yourself on purpose in any way (e.g. by taking an overdose of pills, or by cutting yourself)? | 1. Yes<br>2. No | Self-harm ever<br>1 = "yes"<br>2 = "no" | |

**Table S3. Bullying and Friendship Interview Scale items and coding**

| Bullying item | Response options |
|---|---|
| In the past 6 months: | |
| ***Overt bullying victimisation*** | |
| Someone took personal belongings without asking first | |
| Someone threatened or blackmailed teenager | |
| Someone hit or beat up teenager | |
| Someone played nasty tricks on teenager | 0 – no, has not happened<br>1 – yes, seldom (1-3 times)<br>2 – yes, frequently (>4 times)<br>3 – yes, very frequently (>once a week) |
| Someone called teenager bad/nasty names | |
| ***Relational bullying victimisation*** | |
| Peers would not hang around just to upset teenager | |
| Peers tried to get teenager to do things he/she did not want to do | |
| Peers told lies or nasty stories about teenager | |
| Peers spoilt games to upset teenager | |

**Table S4a. Descriptive statistics and comparison between observed and imputed samples**

| | % missing, (within Imputation sample) | Total Sample | | | Males | | | Females | | |
|---|---|---|---|---|---|---|---|---|---|---|
| | | Imputation dataset (n = 6,934) | Observed (n's vary) | Unobserved (n's vary) | Imputation dataset (n = 3,453) | Observed (n's vary) | Unobserved (n's vary) | Imputation dataset (n = 3,481) | Observed (n's vary) | Unobserved (n's vary) |
| *Exposure* | | | | | | | | | | |
| Emotional Dysregulation | 14.9% | 1.60 (0.013) | 1.58 (0.013) | 1.69 (0.041) | 1.69 (0.018) | 1.68 (0.019) | 1.73 (0.058) | 1.52 (0.018) | 1.49 (0.018) | 1.65 (0.057) |
| *Mediators* | | | | | | | | | | |
| Social Cognition | 17.2% | 2.76 (0.045) | 2.70 (0.047) | 3.06 (0.137) | 3.16 (0.065) | 3.10 (0.074) | 3.44 (0.219) | 2.36 (0.062) | 2.29 (0.056) | 2.70 (0.162) |
| Emotion Recognition | 9.6% | 4.60 (0.034) | 4.60 (0.034) | 4.58 (0.140) | 4.80 (0.048) | 4.81 (0.050) | 4.74 (0.207) | 4.40 (0.048) | 4.40 (0.047) | 4.42 (0.192) |
| Bully Victimisation | 21.4% | 1.81 (0.036) | 1.79 (0.036) | 1.87 (0.102) | 1.78 (0.051) | 1.78 (0.053) | 1.81 (0.139) | 1.84 (0.049) | 1.81 (0.049) | 1.95 (0.142) |
| *Outcomes* | | | | | | | | | | |
| Disordered Eating | 46.1% | 21.7% (0.68) | 23.1% (0.69) | 20.2% (1.23) | 9.4% (0.80) | 8.1% (0.69) | 10.5% (1.34) | 33.9% (1.04) | 33.6% (1.01) | 34.4% (2.22) |
| Self-Harm | 45.0% | 10.4% (0.49) | 10.9% (0.51) | 9.8% (0.91) | 5.6% (0.60) | 5.4% (0.57) | 5.7% (1.00) | 15.3% (0.76) | 14.8% (0.75) | 16.0% (1.63) |

**Table S4b. Descriptive statistics and comparison between observed and imputed samples**

| | % missing, (within Imputation sample) | Total Sample | | | Males | | | Females | | |
|---|---|---|---|---|---|---|---|---|---|---|
| | | Imputation dataset (n = 6,934) | Observed (n's vary) | Unobserved (n's vary) | Imputation dataset (n = 3,453) | Observed (n's vary) | Unobserved (n's vary) | Imputation dataset (n = 3,481) | Observed (n's vary) | Unobserved (n's vary) |
| *Confounders* | | | | | | | | | | |
| Low BMI | 22.3% | 8.2% (0.35) | 8.3% (0.37) | 8.1% (0.88) | 7.8% (0.49) | 7.5% (0.51) | 8.7% (1.27) | 8.6% (0.50) | 9.0% (0.54) | 7.3% (1.24) |
| High BMI | 22.3% | 21.4% (0.52) | 20.8% (0.55) | 23.1% (1.33) | 19.9% (0.72) | 20.1% (0.78) | 19.0% (1.72) | 22.8% (0.76) | 21.5% (0.78) | 27.8% (2.07) |
| Father absence | 15.2% | 21.5% (0.51) | 22.6% (0.55) | 15.4% (1.43) | 21.5% (0.72) | | | 21.5% (0.73) | | |
| Non-white ethnicity | 5.1% | 4.1% (0.25) | 3.9% (0.24) | 7.7% (2.09) | 4.2% (0.36) | | | 4.0% (0.35) | | |
| Maternal eating disorder | 3.3% | 3.6% (0.22) | 3.4% (0.22) | 3.8% (1.95) | 3.6% (0.32) | | | 3.4% (0.31) | | |
| Low income | 11.6% | 15.8% (0.46) | 15.3% (0.46) | 20.2% (1.88) | 16.2% (0.66) | | | 15.5% (0.65) | | |
| Low social class | 7.6% | 4.4% (0.26) | 4.1% (0.25) | 7.7% (1.68) | 3.8% (0.35) | | | 4.9% (0.39) | | |
| Low maternal education | 3.5% | 22.0% (0.51) | 21.7% (0.50) | 31.9% (3.99) | 22.2% (0.72) | | | 21.9% (0.71) | | |
| IQ | 0% | 104.2 (0.20) | | | 104.3 (0.28) | | | 104.1 (0.28) | | |
| Maternal age at delivery | 0% | 29.1 (0.055) | | | 29.3 (0.078) | | | 29.0 (0.077) | | |
| Depression (antenatal) | 6.0% | 6.71 (0.060) | 6.66 (0.060) | 7.48 (0.315) | 6.69 (0.085) | | | 6.72 (0.085) | | |
| Depression (postnatal) | 10.6% | 5.53 (0.058) | 5.48 (0.059) | 5.99 (0.234) | 5.57 (0.082) | | | 5.49 (0.082) | | |

**Table S5. Complete case and imputed association between childhood emotional dysregulation and disordered eating and self-harm in adolescence**

| | Outcome | | Test for main effects [a] | Test for difference between DE and SH estimates [b] | Test for sex-moderation [c] |
|---|---|---|---|---|---|
| | Disordered Eating | Self-Harm | | | |
| *Analysis on Complete Cases (n = 2,846)* | | | | | |
| Unadjusted effect of Emotional Dysregulation | 0.071 [0.019, 0.123] | 0.090 [0.028, 0.151] | 11.46, 0.003 | p = 0.576 | |
| Adjusted for sex | 0.119 [0.063, 0.175] | 0.113 [0.050, 0.176] | 22.81, < 0.001 | p = 0.864 | p = 0.873 |
| Additionally adjusted for confounders C1 | 0.115 [0.057, 0.173] | 0.107 [0.043, 0.172] | 19.75, < 0.001 | p = 0.841 | p = 0.826 |
| Additionally adjusted for confounders C2 | 0.106 [0.046, 0.166] | 0.091 [0.023, 0.158] | 14.57, < 0.001 | p = 0.689 | p = 0.808 |
| Additionally adjusted for confounders C3 | 0.113 [0.053, 0.174] | 0.109 [0.041, 0.178] | 17.56, < 0.001 | p = 0.918 | p = 0.800 |
| *Analysis following Multiple Imputation (n = 6,934)* | | | | | |
| Unadjusted effect of Emotional Dysregulation | 0.048 [0.003, 0.094] | 0.061 [0.011, 0.111] | 7.91, 0.0192 | p = 0.673 | |
| Adjusted for sex | 0.095 [0.045, 0.145] | 0.086 [0.034, 0.138] | 20.10, < 0.001 | p = 0.792 | p = 0.625 |
| Additionally adjusted for confounders C1 | 0.086 [0.036, 0.136] | 0.086 [0.033, 0.140] | 17.57, < 0.001 | p = 1.000 | p = 0.613 |
| Additionally adjusted for confounders C2 | 0.076 [0.024, 0.127] | 0.075 [0.019, 0.131] | 12.47, 0.002 | p = 0.987 | p = 0.602 |
| Additionally adjusted for confounders C3 | 0.082 [0.029, 0.134] | 0.093 [0.036, 0.150] | 15.95, < 0.001 | p = 0.745 | p = 0.600 |

a. Values shown are Wald statistics (2 d.f.) with accompanying p-value
b. P-values for 1 d.f. test based on Wald statistic
c. P-values for 2 d.f. test based on Wald statistic

C1 – parental social class, maternal education, parental income and maternal age at deliver
C2 – maternal depressive symptoms (antenatal and postnatal), maternal history of eating disorder
C3 – child general cognitive ability

**Table S6. Associations between mediators (social cognition, emotion recognition, bullying victimisation) and outcomes (disordered eating, self-harm)**

| | | Males | | | | Females | | | |
|---|---|---|---|---|---|---|---|---|---|
| | | Disordered Eating | | Self-Harm | | Disordered Eating | | Self-Harm | |
| Exposure | | Estimate (SE) | p | Estimate (SE) | p | Estimate (SE) | p | Estimate (SE) | p |
| Social Cognition | Unadjusted | 0.091 (0.046) | 0.051 | 0.043 (0.046) | 0.354 | 0.097 (0.029) | 0.001 | 0.077 (0.032) | 0.019 |
| | Confounder adjusted | 0.047 (0.058) | 0.421 | 0.021 (0.058) | 0.712 | 0.074 (0.035) | 0.032 | 0.027 (0.040) | 0.496 |
| Emotion Recognition | Unadjusted | -0.083 (0.050) | 0.097 | 0.027 (0.050) | 0.594 | -0.029 (0.030) | 0.338 | 0.042 (0.033) | 0.205 |
| | Confounder adjusted | -0.101 (0.050) | 0.045 | 0.044 (0.052) | 0.397 | -0.029 (0.030) | 0.346 | 0.053 (0.034) | 0.124 |
| Bullying Victimisation | Unadjusted | 0.095 (0.041) | 0.021 | 0.055 (0.050) | 0.275 | 0.162 (0.028) | <0.001 | 0.151 (0.032) | <0.001 |
| | Confounder adjusted | 0.083 (0.041) | 0.046 | 0.039 (0.051) | 0.440 | 0.147 (0.028) | <0.001 | 0.056 (0.036) | <0.001 |
| Low BMI | Unadjusted | 0.013 (0.192) | 0.947 | 0.042 (0.178) | 0.812 | -0.453 (0.109) | <0.001 | 0.068 (0.116) | 0.560 |
| | Confounder adjusted | 0.044 (0.203) | 0.829 | 0.117 (0.191) | 0.540 | -0.433 (0.111) | <0.001 | 0.100 (0.120) | 0.406 |
| High BMI | Unadjusted | 0.498 (0.104) | <0.001 | 0.161 (0.122) | 0.188 | 0.325 (0.065) | <0.001 | 0.308 (0.076) | <0.001 |
| | Confounder adjusted | 0.510 (0.109) | <0.001 | 0.219 (0.133) | 0.099 | 0.311 (0.067) | <0.001 | 0.284 (0.079) | <0.001 |

Appropriate confounders for each model are given by the mediation model DAG. For example, Emotional Dysregulation is a confounder for the effect of Social Cognition on Disordered Eating.

Estimates involving continuous exposure and/or outcome have been standardized to enable parameter magnitudes to be compared.

**Table S7. Associations between exposure (emotional dysregulation) and mediators (social cognition, emotion recognition and bullying victimisation)**

| | | | Social Cognition | | Emotion Recognition | | Bullying Victimisation | |
|---|---|---|---|---|---|---|---|---|
| | Exposure | | Estimate (SE) | p | Estimate (SE) | p | Estimate (SE) | p |
| Males | Emotional Dysregulation | Unadjusted | 0.551 (0.014) | <0.001 | 0.094 (0.018) | <0.001 | 0.115 (0.020) | <0.001 |
| | | Confounder adjusted | 0.535 (0.015) | <0.001 | 0.060 (0.020) | 0.002 | 0.124 (0.021) | <0.001 |
| Females | Emotional Dysregulation | Unadjusted | 0.525 (0.015) | <0.001 | 0.071 (0.019) | <0.001 | 0.125 (0.020) | <0.001 |
| | | Confounder adjusted | 0.507 (0.017) | <0.001 | 0.026 (0.020) | 0.195 | 0.104 (0.022) | <0.001 |

Appropriate confounders for each model are given by the mediation model DAG.

Estimates involving continuous exposure and/or outcome have been standardized to enable parameter magnitudes to be compared.

**Figure S1. Flowchart of attrition**

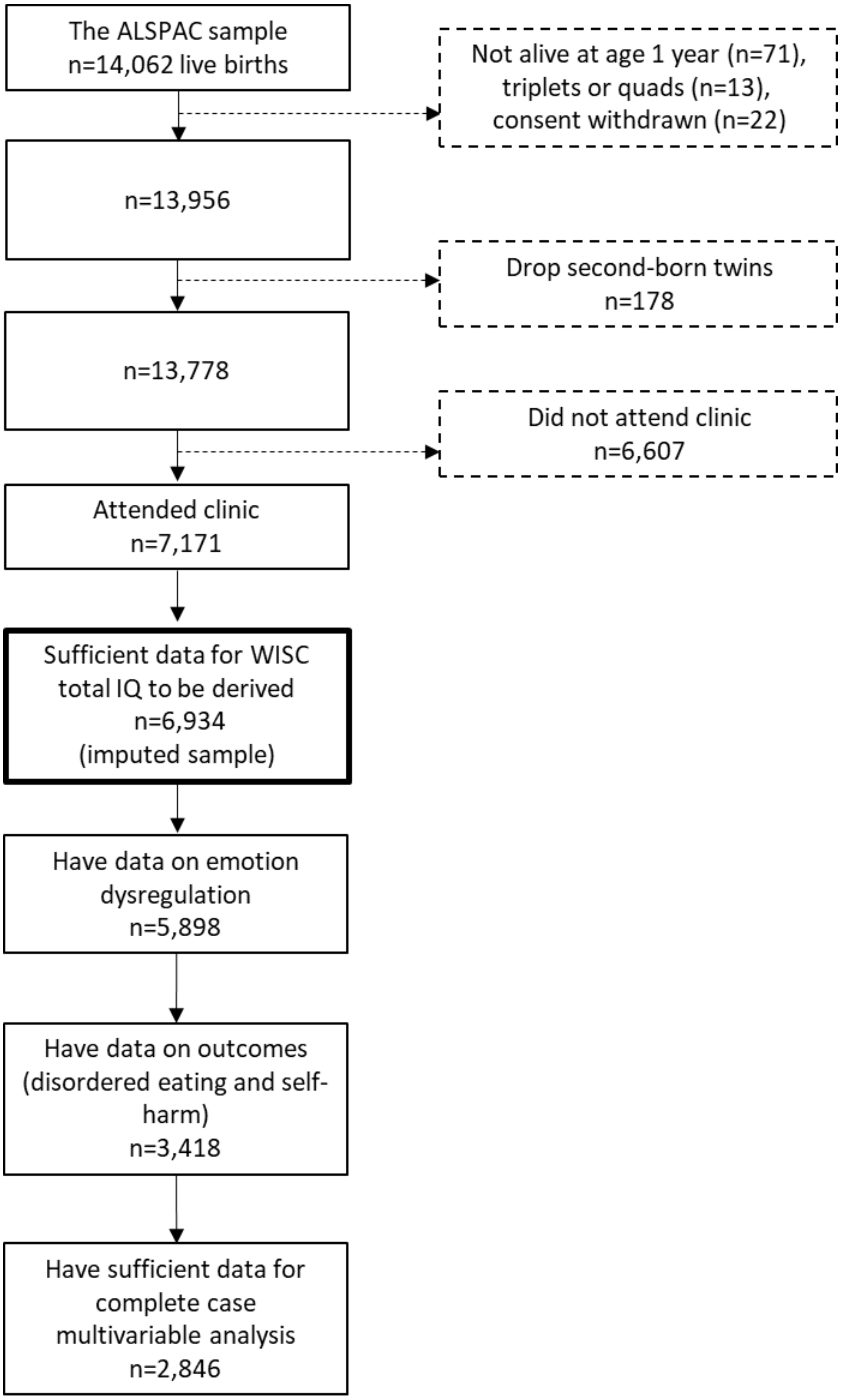

**Figure S2. Partitioning of indirect effects through BMI**

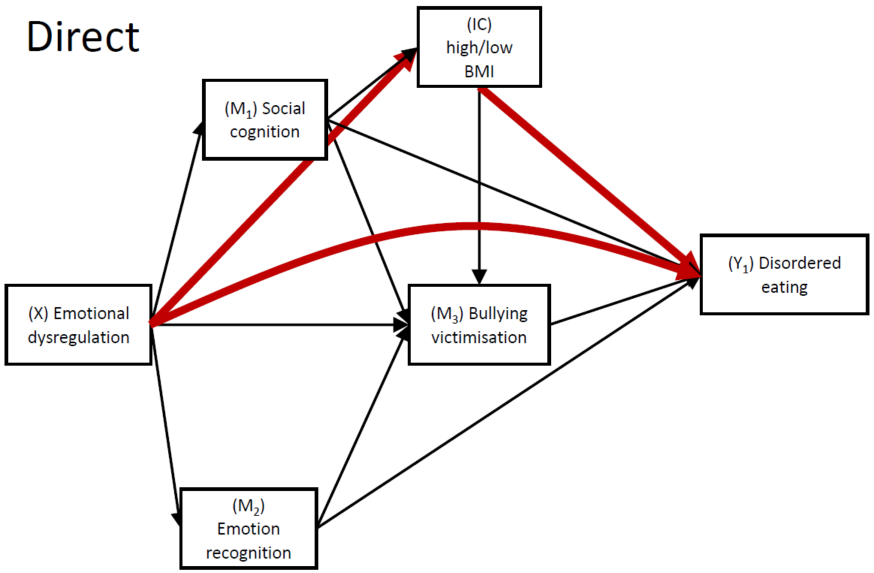


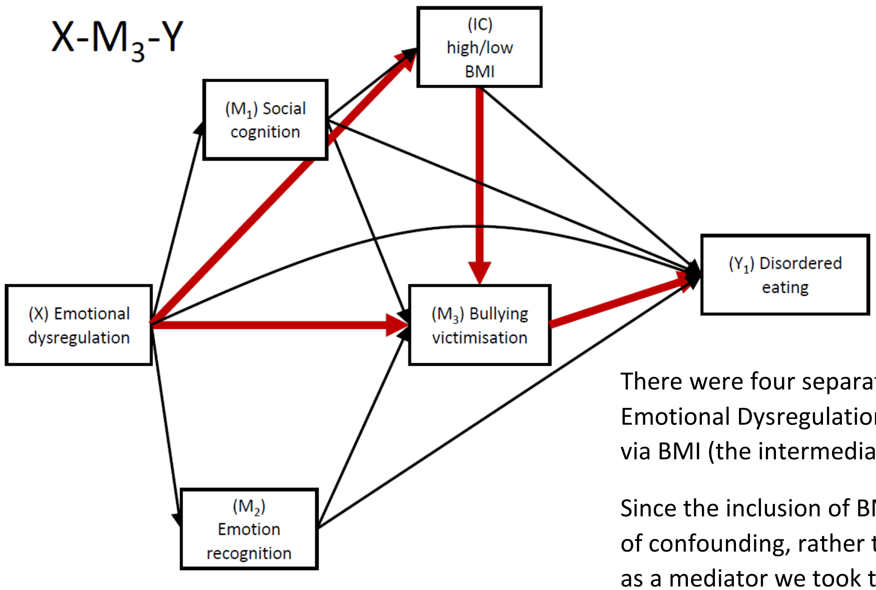


There were four separate indirect paths from Emotional Dysregulation to each outcome that went via BMI (the intermediate confounder).

Since the inclusion of BMI was to address a problem of confounding, rather than examine the role of BMI as a mediator we took the effects for the 4 BMI-paths and combined them with paths that were of substantive interest.

So, for example, the path from X-IC-$Y_1$ (cell 1) can be considered *direct* as far as $M_1$, $M_2$ and $M_3$ are concerned hence the two paths shown here in red were added to yield the direct effect of interest.

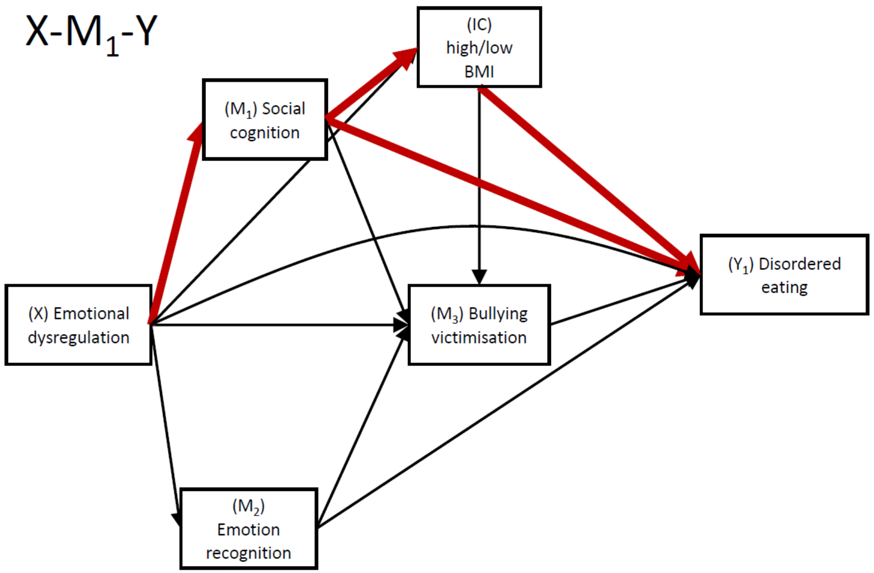


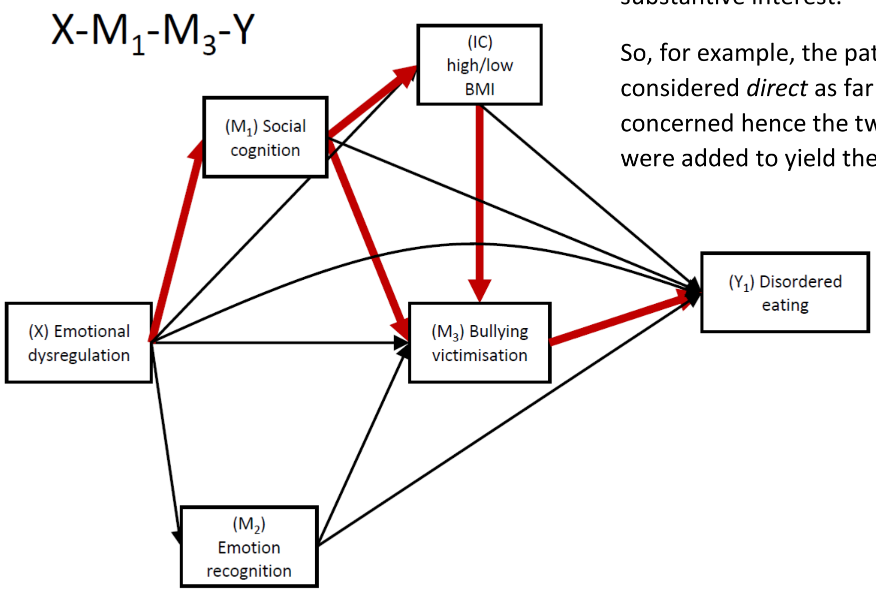

***Appendix References***

Amiri, S., & Behnezhad, S. (2018, October 1). Body mass index and risk of suicide: A systematic review and meta-analysis. *Journal of Affective Disorders*. Elsevier B.V. https://doi.org/10.1016/j.jad.2018.05.028

Anderson, S. E., Sacker, A., Whitaker, R. C., & Kelly, Y. (2017). Self-regulation and household routines at age three and obesity at age eleven: Longitudinal analysis of the UK Millennium Cohort Study. *International Journal of Obesity*, *41*(10), 1459–1466. https://doi.org/10.1038/ijo.2017.94

Andric, S., Maric, N. P., Mihaljevic, M., Mirjanic, T., & van Os, J. (2016). Familial covariation of facial emotion recognition and IQ in schizophrenia. *Psychiatry Research*, *246*, 52–57. https://doi.org/10.1016/j.psychres.2016.09.022

APA. (2013). *Diagnostic and statistical manual of mental disorders* (5th ed). Washington, DC: American Psychiatric Association.

Bould, H., Koupil, I., Dalman, C., Destavola, B., Lewis, G., & Magnusson, C. (2015). Parental mental illness and eating disorders in offspring. *International Journal of Eating Disorders*, *48*(4), 383–391. https://doi.org/10.1002/eat.22325

Bowes, L., Joinson, C., Wolke, D., & Lewis, G. (2015). Peer victimisation during adolescence and its impact on depression in early adulthood: Prospective cohort study in the United Kingdom. *BMJ*, *350*, h2469.

Buitelaar, J. K., Wees, M. van der, Swaab-Barneveld, H., & Gaag, R. J. van der. (1999). Verbal Memory and Performance IQ Predict Theory of Mind and Emotion Recognition Ability in Children with Autistic Spectrum Disorders and in Psychiatric Control Children. *Journal of Child Psychology and Psychiatry*, *40*(6), 869–881. https://doi.org/10.1111/1469-7610.00505

Chang, S. Sen, Chen, Y. Y., Heron, J., Kidger, J., Lewis, G., & Gunnell, D. (2014). IQ and adolescent self-harm behaviours in the ALSPAC birth cohort. *Journal of Affective Disorders*, *152–154*(1), 175–182. https://doi.org/10.1016/j.jad.2013.09.005

Chen, E., & Miller, G. E. (2014). Early-life socioeconomic status, emotion regulation, and the biological mechanisms of disease across the lifespan. In *Handbook of emotion regulation, 2nd ed.* (pp. 586–595). New York, NY, US: The Guilford Press.

Cox, J. L., Holden, J. M., & Sagovsky, R. (1987). Detection of Postnatal Depression: Development of the 10-item Edinburgh Postnatal Depression scale. *British Journal of Psychiatry*, *150*(JUNE), 782–786. https://doi.org/10.1192/bjp.150.6.782

Gianini, L. M., White, M. A., & Masheb, R. M. (2013). Eating pathology, emotion regulation, and emotional overeating in obese adults with binge eating disorder. *Eating Behaviors*, *14*(3), 309–313. https://doi.org/10.1016/j.eatbeh.2013.05.008

Hammami, N., Chaurasia, A., Bigelow, P., & Leatherdale, S. T. (2020). Exploring gender differences in the longitudinal association between bullying and risk behaviours with Body Mass Index among COMPASS youth in Canada. *Preventive Medicine*, *139*, 106188. https://doi.org/10.1016/j.ypmed.2020.106188

Hu, N., Li, J., Glauert, R. A., & Taylor, C. L. (2017). Influence of exposure to perinatal risk factors and parental mental health related hospital admission on adolescent deliberate self-harm risk. *European Child and Adolescent Psychiatry*, *26*(7), 791–803. https://doi.org/10.1007/s00787-017-0948-4

Hughes, R. A., Heron, J., Sterne, J. A. C., & Tilling, K. (2019). Accounting for missing data in statistical analyses: Multiple imputation is not always the answer. *International Journal of Epidemiology*, *48*(4), 1294–1304.

Janssen, I., Craig, W. M., Boyce, W. F., & Pickett, W. (2004). Associations between Overweight and Obesity with Bullying Behaviors in School-Aged Children. *Pediatrics*, *113*(5 I), 1187–1194. https://doi.org/10.1542/peds.113.5.1187

Kinoshita, K., Kinoshita, Y., Shimodera, S., Nishida, A., Inoue, K., Watanabe, N., … Okazaki, Y. (2012). Not only body weight perception but also body mass index is relevant to suicidal ideation and self-harming behavior in Japanese adolescents. *Journal of Nervous and Mental Disease*, *200*(4), 305–309. https://doi.org/10.1097/NMD.0b013e31824cb29b

Lodebo, B. T., Möller, J., Larsson, J. O., & Engström, K. (2017). Socioeconomic position and self-harm among adolescents: A population-based cohort study in Stockholm, Sweden. *Child and Adolescent Psychiatry and Mental Health*, *11*(1), 46. https://doi.org/10.1186/s13034-017-0184-1

Magnusson, P. K. E., Rasmussen, F., Lawlor, D. A., Tynelius, P., & Gunnell, D. (2006). Association of body mass index with suicide mortality: A prospective cohort study of more than one million men. *American Journal of Epidemiology*, *163*(1), 1–8. https://doi.org/10.1093/aje/kwj002

Mewton, L., Champion, K., Kay-Lambkin, F., Sunderland, M., Thornton, L., & Teesson, M. (2019, January 14). Lifestyle risk indices in adolescence and their relationships to adolescent disease burden: Findings from an Australian national survey. *BMC Public Health*. BioMed Central Ltd. https://doi.org/10.1186/s12889-019-6396-y

Mitchison, D., Hay, P., Slewa-Younan, S., & Mond, J. (2014). The changing demographic profile of eating disorder behaviors in the community. *BMC Public Health*, *14*(1), 1–9. https://doi.org/10.1186/1471-2458-14-943

Nowicki, S., & Duke, M. P. (1994). Individual differences in the nonverbal communication of affect: The diagnostic analysis of nonverbal accuracy scale. *Journal of Nonverbal Behavior*, *18*(1), 9–35.

Nyberg, J., Gustavsson, S., Åberg, M. A. I., Kuhn, H. G., & Waern, M. (2019). Late-adolescent risk factors for suicide and self-harm in middle-aged men: explorative prospective population-based study. *The British Journal of Psychiatry*, *217*(1), 1–7. https://doi.org/10.1192/bjp.2019.243

Rai, D., Culpin, I., Heuvelman, H., Magnusson, C. M. K., Carpenter, P., Jones, H. J., … Pearson, R. M. (2018). Association of autistic traits with depression from childhood to age 18 years. *JAMA Psychiatry*, *75*(8), 835–843.

Reed, Z. E., Mahedy, L., Jackson, A., Davey Smith, G., Penton-Voak, I., Attwood, A. S., & Munafò, M. R. (2021). Examining the bidirectional association between emotion recognition and social autistic traits using observational and genetic analyses. *Journal of Child Psychology and Psychiatry*, *62*(11), 1330–1338.

Sainsbury, K., Evans, E. H., Pedersen, S., Marques, M. M., Teixeira, P. J., Lähteenmäki, L., … Sniehotta, F. F. (2019). Attribution of weight regain to emotional reasons amongst European adults with overweight and obesity who regained weight following a weight loss attempt. *Eating and Weight Disorders*, *24*(2), 351–361. https://doi.org/10.1007/s40519-018-0487-0

Skuse, D. H., Mandy, W. P. L., & Scourfield, J. (2005). Measuring autistic traits : heritability , reliability and validity of the Social and Communication Disorders Checklist. *British Journal of Psychiatry*, *187*, 568–572.

StataCorp. (2019). Stata Statistical Software: Release 16. College Station, TX: StataCorp LLC.

Stein, A., Pearson, R. M., Goodman, S. H., Rapa, E., Rahman, A., McCallum, M., … Pariante, C. M. (2014, November 15). Effects of perinatal mental disorders on the fetus and child. *The Lancet*. Lancet Publishing Group. https://doi.org/10.1016/S0140-6736(14)61277-0

Troy, A. S., Ford, B. Q., McRae, K., Zarolia, P., & Mauss, I. B. (2017). Change the things you can: Emotion regulation is more beneficial for people from lower than from higher socioeconomic status. *Emotion*, *17*(1), 141–154. https://doi.org/10.1037/emo0000210

Vidmar, S., Carlin, J., Hesketh, K., & Cole, T. (2013). Standardizing anthropometric measures in children and adolescents with functions for egen: Update. *Stata Journal*, *13*(2), 366–378. https://doi.org/10.1177/1536867x1301300211

Wechsler, D., Golombok, S., & Rust, J. (1992). *WISC-III UK Wechsler Intelligence Scale for Children – Third Edition UK Manual.* The Psychological Corporation.

Wolke, D., Woods, S., Stanford, K., & Schulz, H. (2001). Bullying and victimization of primary school children in England and Germany: Prevalence and school factors. *British Journal of Psychology*, *92*(4), 673–696.
